# A proof-of-concept investigation into predicting follicular carcinoma on ultrasound using topological data analysis and radiomics

**DOI:** 10.1101/2023.10.18.23297210

**Authors:** Andrew M Thomas, Ann C Lin, Grace Deng, Yuchen Xu, Gustavo Fernandez Ranvier, Aida Taye, David S Matteson, Denise Lee

**Affiliations:** Department of Statistics and Actuarial Science, University of Iowa; Department of Surgery, Icahn School of Medicine at Mount Sinai; Department of Statistics and Data Science, Cornell University

**Keywords:** topological data analysis, follicular carcinoma, multimodal, thyroid nodule, machine learning

## Abstract

**Aims:** Sonographic risk patterns identified in established risk stratification systems (RSS) may not accurately stratify follicular carcinoma from adenoma, which share many similar US characteristics. The purpose of this study is to investigate the performance of a multimodal machine learning model utilizing radiomics and topological data analysis (TDA) to predict malignancy in follicular thyroid neoplasms on ultrasound.

**Methods:** This is a retrospective study of patients who underwent thyroidectomy with pathology confirmed follicular adenoma or carcinoma at a single academic medical center between 2010–2022. Features derived from radiomics and TDA were calculated from processed ultrasound images and high-dimensional features in each modality were projected onto their first two principal components. Logistic regression with L2 penalty was used to predict malignancy and performance was evaluated using leave-one-out cross-validation and area under the curve (AUC).

**Results:** Patients with follicular adenomas (n=7) and follicular carcinomas (n=11) with available imaging were included. The best multimodal model achieved an AUC of 0.88 (95% CI: [0.85, 1]), whereas the best radiomics model achieved an AUC of 0.68 (95% CI: [0.61, 0.84]).

**Conclusions:** We demonstrate that inclusion of topological features yields strong improvement over radiomics-based features alone in the prediction of follicular carcinoma on ultrasound. Despite low volume data, the TDA features explicitly capture shape information that likely augments performance of the multimodal machine learning model. This approach suggests that a quantitative based US RSS may contribute to the preoperative prediction of follicular carcinoma.

## Introduction

Thyroid nodules are common in the general population, and standard evaluation begins with ultrasound imaging. The decision to proceed with fine needle aspiration biopsy (FNAB) is based on nodule size and the presence of suspicious sonographic features [1]. Multiple risk stratification systems (RSS), such as the Thyroid Imaging Reporting and Data System (TI-RADS) and American Thyroid Association (ATA) sonographic pattern guidelines, have been developed to systematically evaluate thyroid ultrasounds and to estimate malignancy risk in thyroid nodules [2,3]. However, there continues to be a lack of standardization in the use of these various RSSs for thyroid nodules, as well as in the clinical interpretation of their assigned values [4–6]. Additionally, distinguishing benign follicular adenomas from follicular carcinoma has posed a particular diagnostic dilemma, as they share many similar sonographic characteristics and may not be accurately stratified on RSSs [7,8]. Since follicular thyroid neoplasms cannot be distinguished on cytology, final diagnosis can only be made after surgical excision with microscopic examination for capsular or vascular invasion [9,10]. An estimated 80-90% of follicular thyroid neoplasms are benign, resulting in a significant number of surgeries performed for benign disease [11].

In recent years, the paradigm of work up of thyroid nodules has shifted to a more conservative approach to prevent unnecessary biopsies, and ultimately unnecessary surgeries. As such, the implementation of innovative techniques to improve risk stratification of follicular thyroid neoplasms based solely on non-invasive tools, such as ultrasound, are of particular interest. Quantitative medical imaging analysis is the process of extracting large numbers of high-dimensional textural features from tomographic images, which can then be used to inform decision support tools. Radiomics specifically refers to extraction of pixel-level features within a region of interest of the image, converting the image into high-dimensional data [12]. Alternatively, topological data analysis (TDA) is a field of mathematics that specifically utilizes tools from algebraic topology, such as persistent homology and persistence diagrams [13], to examine structures on multiple scales, focusing on the shape of data [14,15].

While these techniques can be applied to many clinical scenarios, this area of research has had its particular focus in oncology, as these processes have potential to identify and refine tumor phenotypes on imaging beyond the scope of human analysis. Previous studies have applied radiomics to thyroid ultrasound, with promising results in predicting malignancy in thyroid nodules in comparison to radiologist review using RSSs [16,17]. And while TDA has been applied to the diagnosis of melanoma, breast, brain, colorectal, and lung cancer, there is a lack of research in the utilization of TDA in thyroid imaging [14,18].

These two quantitative imaging analysis techniques have potential to enhance the diagnostic ability of ultrasound images in distinguishing follicular thyroid neoplasms by capturing features that are imperceptible to the human eye. The objective of this study is to investigate the performance of a multimodal machine learning (ML) model utilizing both radiomics and TDA to predict malignancy in follicular thyroid neoplasms on ultrasound images. This is a follow-up to a previous proof-of-concept study [19] examining the improvement of multimodal inclusion of radiomics features in a baseline model of clinical variables. The purpose of this study is to investigate the performance of topological features in predicting malignancy using solely thyroid ultrasound images.

## Methods

### Data Collection

This study was approved by the Icahn School of Medicine Institutional Review Board. A cohort of adult patients (N=908) who underwent thyroid surgery at our tertiary referral institution between 2010 and 2022 were identified. Of these, patients with diagnosis of follicular carcinoma or follicular adenoma on final pathology were reviewed (N=82). Exclusion criteria were as follows: history of prior thyroid cancer (N=5), multiple cancer diagnoses in final pathology (N=2), diagnosis of Hurthle cell adenoma or carcinoma (N=7), no pre-operative thyroid ultrasound images available (N=50). A total of 18 patients remained for inclusion in this study, with N=7 patients in follicular adenoma cohort and N=11 in follicular carcinoma.

All available pre-operative thyroid ultrasound images for each patient were downloaded from the Picture Archiving and Communications System (PACS) for analysis. Each nodule of interest was identified by cross-referencing the images with cytopathology and final pathology reports. The images including the nodule in view were selected and annotated to indicate region of interest (ROI) by an endocrine surgery fellow (A.C.L). For every patient, a representative image was chosen, and downscaled to the minimum image resolution so that each image would have a similar number of pixels.

### Topological Data Analysis

Topological data analysis (TDA) has been effectively employed to extract predictive features from medical imaging for classification tasks [20–23]. One of the most commonly employed tools within TDA is persistent homology [13]. Persistent homology is a mathematical tool that can be used to detect quantitative shape information such as connected components (0-dimensional) and holes/loops (1-dimensional) that capture contiguous dark and white regions of an image, respectively. The persistent homology of an image is summarized by its persistence diagram. Useful information corresponding to various shape features from the image can be extracted from the persistence diagram and then enhanced as inputs for machine learning algorithms using filtrations such as the *height filtration*, *greyscale filtration,* and *Vietoris-Rips filtration* [24]. A major advantage of TDA is its ability to capture shape content explicitly [25] and its high performance in small sample sizes [26,27], which is relevant for low incidence populations such as follicular carcinoma. Further information on details of persistence homology calculations for this study can be found in the Supplementary Material.

### Model Predictors

To apply the height filtration to a greyscale image, we must first binarize the image. As it is not obvious which scale, threshold, or direction we should choose to capture the most relevant shape information, we derive a persistence diagram for a variety of different scales, thresholds, and directions. Thus, 180 persistence diagrams in dimensions 0 and 1 were derived for each image by first smoothing the image according to a Gaussian filter at 3 different bandwidths, binarizing the images at 6 different thresholds, then calculating the persistent homology of the height filtration in 5 separate directions. Each of these 180 diagrams was converted into a numerical value by taking its persistent entropy, conveying topological and geometric information for each annotated nodule (*TDA_height*). A total of 910 radiomics features were extracted for each image, similar to methods outlined in a previous study [19]. To reduce the risk of model saturation, the 180 topological features and 910 radiomics features were projected onto their first two principal components (See Supplementary Material). These features were denoted as *TDA_height_pca1*, *TDA_height_pca2*, *Rad_pca1*, *Rad_pca2*. (**Table I**)

**Table I:** Table of every considered model feature and their derivation, along with potential corresponding radiographic features.

| **Model predictor name** | **TDA or Radiomics derived** | **Features prior to PCA reduction** | **Potential Corresponding radiographic feature** |
| --- | --- | --- | --- |
| TDA_height_pca1 | TDA | 180 | Generic shape of nodule |
| TDA_height_pca2 | TDA | 910 | Irregularity of borders |
| Rad_pca1 | Radiomics | NA | Complex image contrast and pixel correlation |
| Rad_pca2 | Radiomics | NA | Complex image contrast and pixel correlation |
| Rad_mean | Radiomics | NA | Image brightness/lightness |
| Rad_variance | Radiomics | NA | Image contrast |
| Rad_ratio | Radiomics | NA | Wider-than-tall or taller-than-wide |
| TDA_alps_grey | TDA | NA | Spongiform (or lack thereof) nature of nodule |
| TDA_alps_boundary | TDA | NA | Generic shape of nodule |

In addition to the above, the following additional radiomic features were obtained: mean and variance of the pixel intensity and aspect ratio of the images (*Rad_mean*; *Rad_variance*; *Rad_ratio*). Two additional topological features derived from the ALPS statistic were also obtained [28]: one feature from the image boundary using the Vietoris-Rips filtration (a proxy for the irregularity of the nodule boundary) and another from the greyscale filtration for the entire image (a proxy for the number of black holes in the image) (*TDA_alps_boundary*, *TDA_alps_grey*, see **Table I**). The correlations between these various features for all representative images can be seen in **Figure 1**.

**Figure 1:**
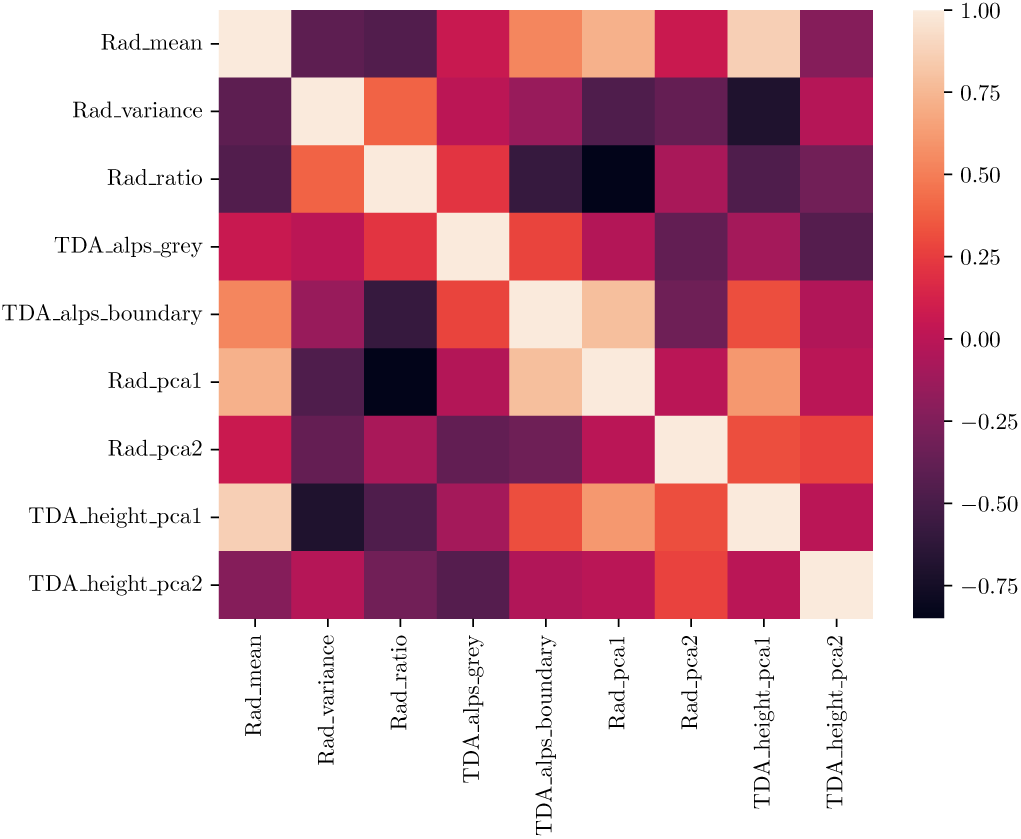
(Pearson) correlations between each of the predictors. One can see that there is some correlation between some of the radiomics features (such as Rad_mean and Rad_pca1) and the TDA features. Ultimately the TDA features produce the better models.

### Model Development

A total of 6 models were considered. The first 3 models consisted of all features of a given type: the complete “All Predictors” model, the “Radiomics only” model (all 5 Radiomics features), the “TDA only” model (all 4 TDA features). An exhaustive search of all 511 possible submodels of our 9 predictor model (2^9^ – 1 = 511) were then evaluated, with a default value of regularization *C* = 1. Of the 511 possible models, the three best performing models were selected from both radiomics and TDA features (“Best Multimodal”), from Radiomics only features (“Best Radiomics”) and from TDA only features (“Best TDA”) (**Table II**). For our analysis we used regularized logistic regression with an *l*^2^ penalty, because it has desirable theoretical properties that lead it to be less likely to overfit [29]. The (unlabeled) dataset was defined as *X* = (*X*_1_,…,*X*_18_). Models were validated using leave-one-out cross-validation to estimate the area under the curve (AUC) of the ROC curve [30]. The predicted probabilities for the ROC curve π_1_,…,π_18_ were defined with π_i_ as the predicted probability of a follicular carcinoma for patient *i*, based on a model trained on all observations besides *i*.

**Table II.** List of predictors included in each of the six main models of the paper.

|  | **All Predictors** | **Radiomics only** | **TDA only** | **Best Multimodal** | **Best Radiomics** | **Best TDA** |
| --- | --- | --- | --- | --- | --- | --- |
| TDA_height_pca1 | **X** |  | **X** | **X** |  |  |
| TDA_height_pca2 | **X** |  | **X** | **X** |  | **X** |
| Rad_pca1 | **X** | **X** |  |  |  |  |
| Rad_pca2 | **X** | **X** |  |  |  |  |
| Rad_mean | **X** | **X** |  | **X** |  |  |
| Rad_variance | **X** | **X** |  |  |  |  |
| Rad_ratio | **X** | **X** |  |  | **X** |  |
| TDA_alps_grey | **X** |  | **X** |  |  |  |
| TDA_alps_boundary | **X** |  | **X** | **X** |  | **X** |

## Results

### Evaluation of Model Performance

The best performing model comprising both TDA and Radiomics features achieved an AUC of 0.88, a sensitivity of 0.91, and a specificity of 0.57 (for a threshold of 0.5). This was denoted as the “Best Multimodal” model. The best model with only Radiomics features included yielded an AUC of 0.68, a sensitivity of 0.91, and a specificity of 0.43. This was denoted as the “Best Radiomics” model. The best model with only TDA features was denoted as the “Best TDA” and had an AUC of 0.88 (sensitivity 0.91, specificity 0.71). Corresponding AUCs for the “All Predictors”, “TDA only”, and “Radiomics only” models were 0.74 (sensitivity 0.91, specificity 0.43), 0.82 (sensitivity 0.91, specificity 0.71), and 0.49 (sensitivity 0.73, specificity 0.43) respectively.

For most models except “Best Multimodal” the optimal amount of regularization was around 1 (**Figure 2**), demonstrating the “Best Multimodal” model’s strength by choosing λ such that its maximum AUC was not achieved. Therefore, the regularization parameter was fixed at its original value λ = 1. Given that λ tended towards 0 from its moderate values—meaning overfitting or large parameter values were not penalized—the Best Multimodal, All Predictors, and Best Radiomics models did not decline in terms of AUC. The other models exhibited a sort of unimodal structure to these model selection curves, suggesting that the multimodal model is best at capturing *legitimate* associations within the dataset, though the “Best TDA” performed just about as well. Nodules along with their actual status and predicted probability for the “Best Multimodal” model, can be seen in **Figure 3**.

**Figure 2:**
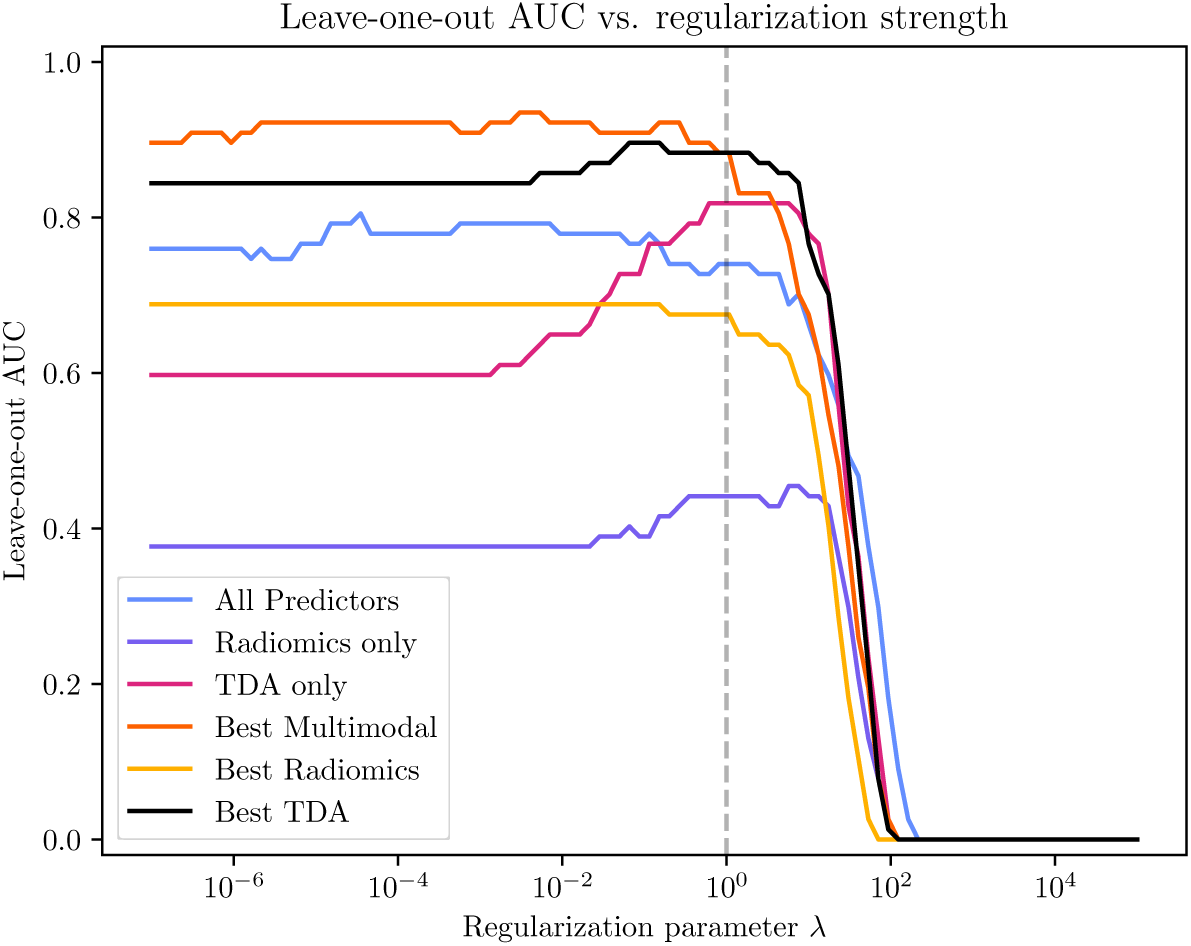
Plot of leave-one-out AUC vs. regularization strength for each of the six main models considered in this paper. Vertical line indicates where regularization parameter equals 1.

**Figure 3:**
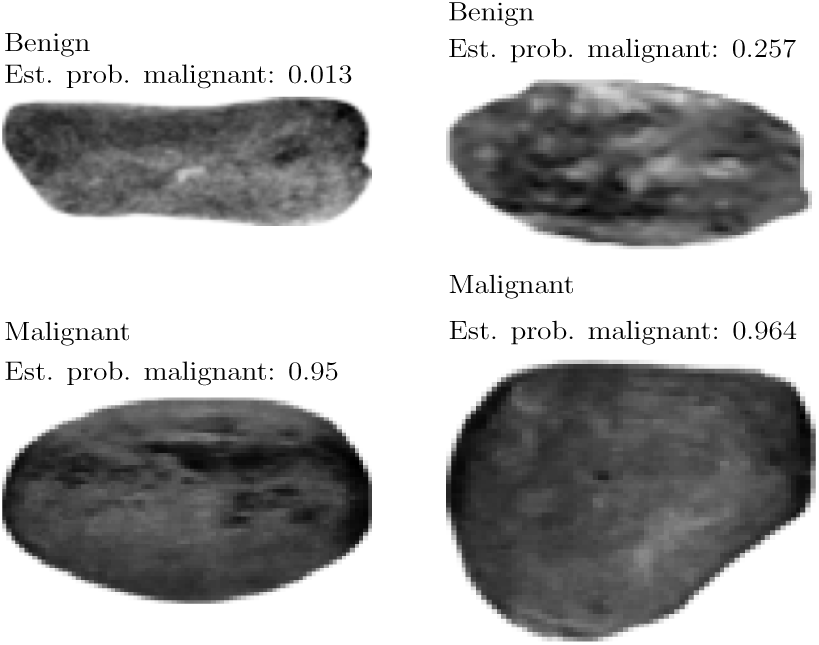
Representative nodules for lowest and highest predicted probabilities of malignancy respectively under the “Best Multimodal” model along with true classification.

### Model Validation

The subsampling bootstrap was used to evaluate the uncertainty of each AUC, sensitivity, and specificity for the five models [31]. Sensitivity and specificity were calculated with prediction threshold 0.5. To estimate the sampling distribution of each metric, 1000 random samples of size *b =* 10 were uniformly chosen at random from the set of all subsets where 4 patients had adenomas and 6 had carcinomas, and then each metric was evaluated on said sample. This approach, as opposed to a naïve bootstrap method, ensured that no duplicate observations appeared in our example, so that the bootstrap samples did not have the opportunity to train on examples it would be tested on. Unlike a conventional asymptotic confidence interval, the values from which these intervals were calculated can be considered as from the correct image distribution. The values of Leave-one-out AUC, sensitivity, and specificity for each of the 5 main models along with their 95% confidence intervals can be seen in **Figures 4–6** and **Table III**. Confidence intervals for the AUCs using DeLong’s test [32] are listed in **Table IV**.

**Figure 4:**
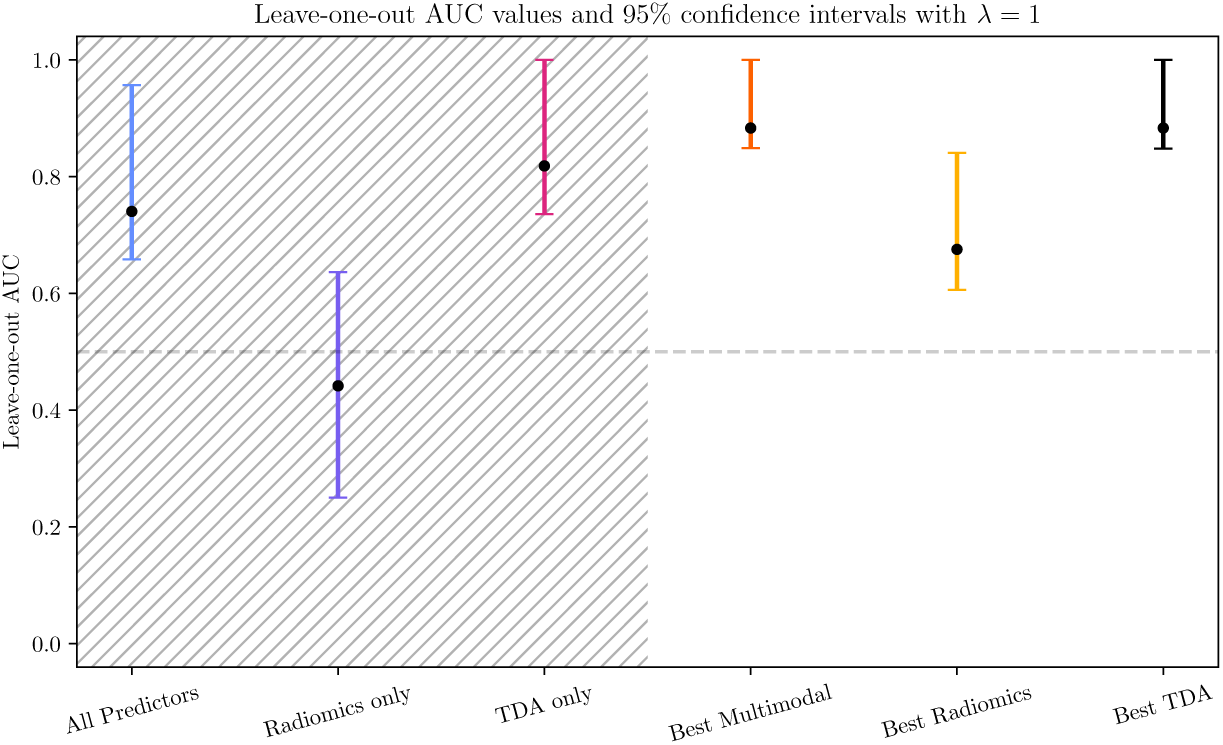
95% subsampling bootstrap (marginal) confidence intervals for the AUC of each of the six main models considered in this paper. Shaded region corresponds to “full” models; unshaded region corresponds to the best models after model selection based on leave-one-out AUC.

**Figure 5:**
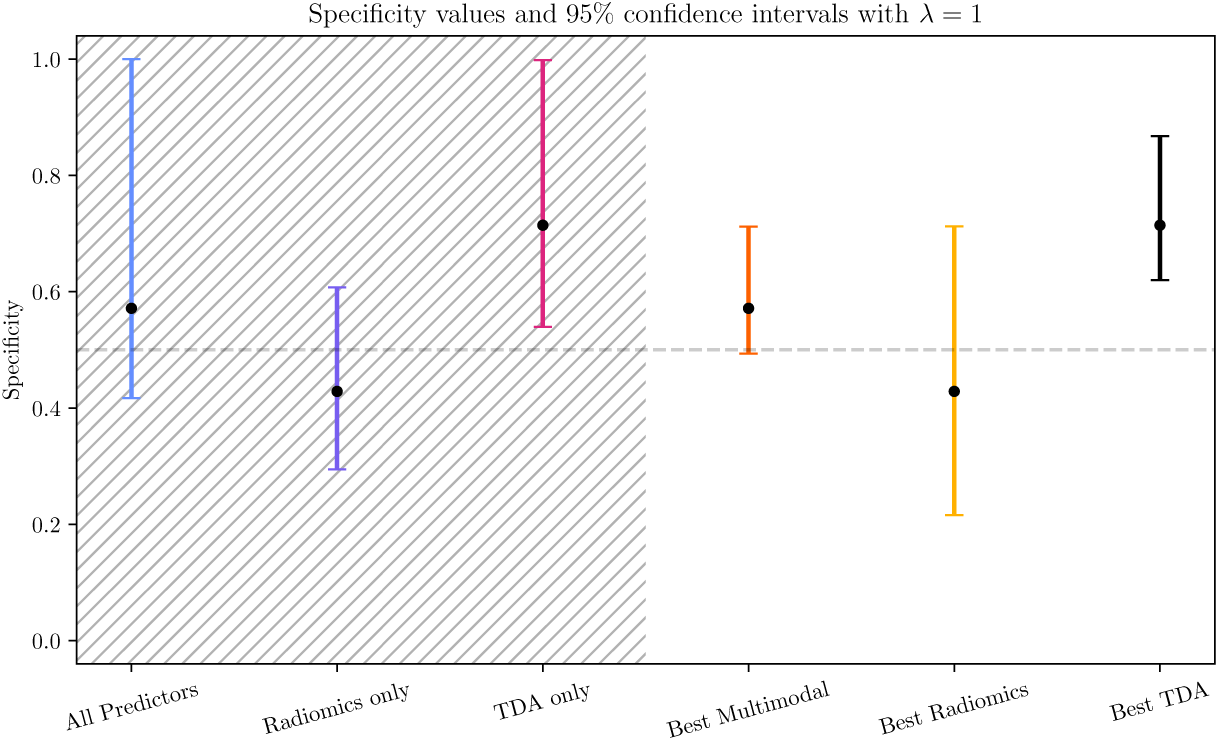
95% subsampling bootstrap (marginal) confidence intervals for the sensitivity of each of the six main models considered in this paper. Shaded region corresponds to “full” models; unshaded region corresponds to the best models after model selection based on leave-one-out AUC.

**Figure 6:**
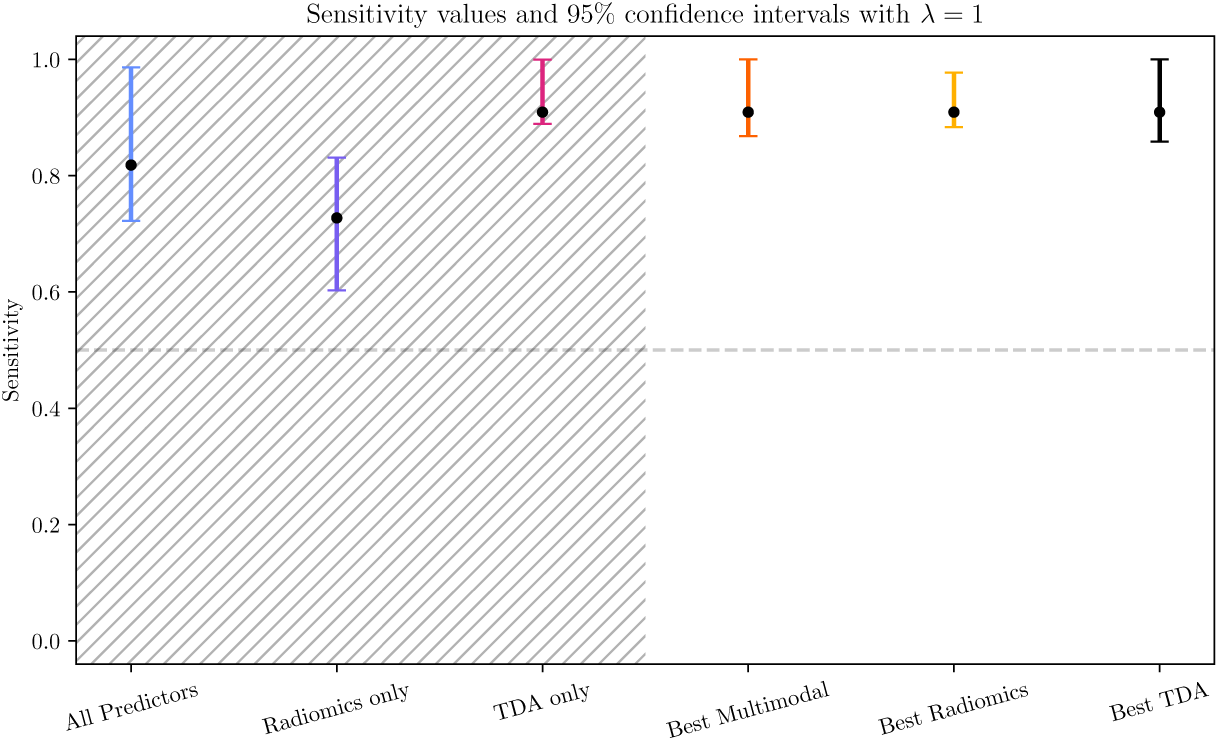
95% subsampling bootstrap (marginal) confidence intervals for the specificity of each of the six main models considered in this paper. Shaded region corresponds to “full” models; unshaded region corresponds to the best models after model selection based on leave-one-out AUC.

**Table III.**
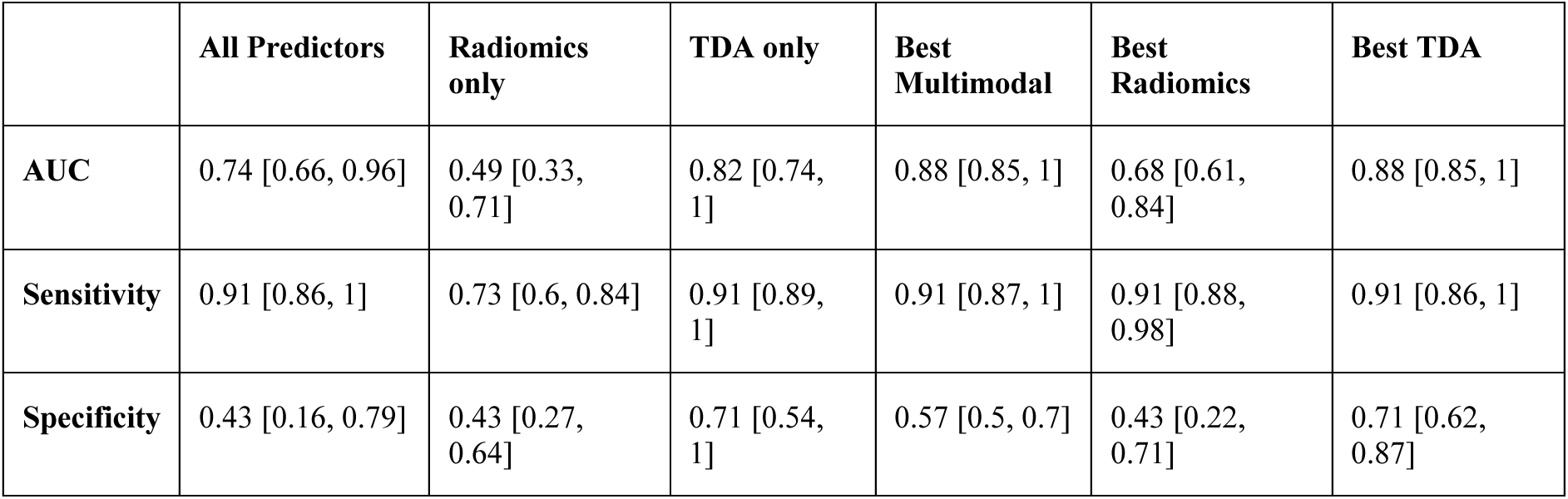
95% bootstrap (marginal) confidence intervals for the AUC, sensitivity, and specificity of each of the five main models considered in this paper, as well as seen in Figures 2–4.

|  | **All Predictors** | **Radiomics only** | **TDA only** | **Best Multimodal** | **Best Radiomics** | **Best TDA** |
| --- | --- | --- | --- | --- | --- | --- |
| **AUC** | 0.74 [0.66, 0.96] | 0.49 [0.33, 0.71] | 0.82 [0.74, 1] | 0.88 [0.85, 1] | 0.68 [0.61, 0.84] | 0.88 [0.85, 1] |
| **Sensitivity** | 0.91 [0.86, 1] | 0.73 [0.6, 0.84] | 0.91 [0.89, 1] | 0.91 [0.87, 1] | 0.91 [0.88, 0.98] | 0.91 [0.86, 1] |
| **Specificity** | 0.43 [0.16, 0.79] | 0.43 [0.27, 0.64] | 0.71 [0.54, 1] | 0.57 [0.5, 0.7] | 0.43 [0.22, 0.71] | 0.71 [0.62, 0.87] |

**Table IV.** 95% asymptotic (marginal) confidence intervals for AUC using DeLong’s test.

|  | **All Predictors** | **Radiomics only** | **TDA only** | **Best Multimodal** | **Best Radiomics** | **Best TDA** |
| --- | --- | --- | --- | --- | --- | --- |
| **AUC** | 0.74 [0.49, 1] | 0.49 [0.12, 0.76] | 0.82 [0.59, 1] | 0.88 [0.72, 1] | 0.68 [0.38, 0.97] | 0.88 [0.71, 1] |

## Discussion

We developed a multimodal ML classifier model incorporating both TDA and radiomics to generate predictions of carcinoma versus adenoma from ultrasound images of thyroid follicular lesions. Our “Best Multimodal” model achieved an AUC of 0.88, outperforming the models that included only radiomics features and only TDA features. The “Best TDA” model consisted of only two predictors and performed as well as the “Best Multimodal” model (AUC = 0.88). The presence of only two predictors in the “Best TDA” model strongly suggests that topological features are capturing genuine shape information that distinguish follicular carcinoma from adenoma. To the best of our knowledge, this is the first study to apply TDA to distinguish follicular carcinoma from adenoma using thyroid ultrasound images and to demonstrate the potential of topological features to not only augment the predictive power of ML classifier models, but also capture legitimate distinctions between follicular carcinoma and adenoma.

Our ability to evaluate thyroid nodules using ultrasound is currently limited by the lack of a standardized approach to manual interpretation as well as significant interobserver variability. As a means to improve the diagnostic power of thyroid ultrasound, quantitative imaging techniques have been studied to automate evaluation of thyroid nodules. TDA is especially amenable to extracting specific features within the “ATA Nodule Sonography Pattern Risk of Malignancy” [3], which can be described topologically. For example in the greyscale filtration, a cyst would show low (0-dimensional) persistent entropy as it consists of a single, uniform dark region; a spongiform nodule would contain many significant 0-dimensional connected components in its persistence diagram. Microcalcifications would present as many significant holes in the 1-dimensional greyscale filtration persistence diagram. As previously mentioned, irregular margins (via notions of fractal dimension [33]) and generic shape features (such as an acute angle interface, using the height filtration) [25] can be captured topologically. With a larger dataset (best practices suggest 10-15 samples per radiomics feature [34]) a combination of topological and radiomics features may enhance the ability to capture inherent and distinct nodule characteristics as many traditional radiomics features can improve interpretability and capture relevant pixel statistics that TDA features capture only indirectly (such as statistics relating to contrast).

We selected topological features known throughout the literature to either correspond to relevant nodule patterns or capture shape effectively. Submodels were then chosen according to the inclusion of the various radiomics and TDA features or by model performance, evaluated by AUC (**Figures 2 and 4)**. The “Radiomics only” model had the poorest performance of all six models evaluated, with an AUC of 0.5, which is equivalent to random guessing. Additionally, there is minimal overlap of the confidence intervals for the AUCs of the Best Multimodal/Best TDA (0.88) and Best Radiomics models (0.68), suggesting that with additional data a multimodal model including TDA features may be *statistically significantly* better than a model containing only Radiomics features to classify follicular carcinoma on ultrasound.

A strength of this paper is that few studies using ML have focused specifically on classifying follicular subtypes in thyroid nodules. Shin et al investigated the use of radiomics alone to develop a ML classifier model to differentiate between follicular adenoma and carcinoma using ultrasound images [17]. Our prior proof-of-concept study developed a multimodal ML model that incorporated both clinical data and radiomics to predict follicular adenoma from carcinoma. Yu et al combined radiomics features with clinical variables to distinguish follicular neoplasms on ultrasound [35]. Compared to the results of these papers, our addition of TDA features seem to improve the classification of follicular neoplasms on ultrasound. Our multimodal TDA+Radiomics image-only method performed better than the image-only models in Shin et al (maximum leave-one-out AUC of 0.646, 95% CI: 0.544–0.653). Our prior study demonstrated only a modest improvement in predictive power using both clinical and radiomics data over radiomics or clinical only data [19]. The radiomics image-only model of Yu et al, which also used logistic regression, achieved an AUC of 0.772 (95% CI: 0.707–0.838) . For the deep learning model of Seo et al [36], the authors used a convolutional neural network trained on nodule boundary subimages and predicted nodule status by majority vote. The method of Seo et al achieved a higher performance on image features alone than the studies of Shin et al and Yu et al (AUC of 0.809 on test set). Though our study suggests TDA might enhance prediction of follicular carcinoma from adenoma on ultrasound imaging, additional studies will be required to establish the robustness of these preliminary findings. Given that follicular thyroid neoplasms pose a particular diagnostic dilemma within thyroid pathology, the ability of innovative techniques such as TDA to enhance pre-operative classification of these lesions and subsequently prevent unnecessary surgeries will continue to be an area of focus.

In the field of medical imaging analysis, TDA has been previously applied to computed tomography (CT) and magnetic resonance imaging (MRI), in an attempt to capture and extract information from these images to allow for automated diagnosis or stratification [37]. Specifically, previous studies have looked to utilize TDA summaries of medical images to stratify COPD, detect COVID-19 infection, and detect osteoarthritis in earlier stages, showing potential for better performance over manual as well as gold standard machine learning models [38–40]. In a similar setting to this one, TDA, Radiomics, and multimodal models were contrasted in the context of lung tumor histology prediction [18]. However, there is still significant work to be done in validating, improving access to, and encouraging the widespread use of TDA.

Our study contained several limitations. First, our study included a small number of patients, and it will be important for future studies to use large multicenter datasets to develop models that are generalizable to the larger population. Second, our patient population often are referred with ultrasound images from different institutions and machine types, and thus, there was significant variability in the quality of the images. Additionally, we did not include other follicular pattern thyroid lesions, such as non-invasive follicular thyroid neoplasm with papillary-like nuclear features (NIFTP) or follicular variant papillary thyroid cancer. Third, we did not perform comparison of our model’s performance to any currently available radiologic RSS (i.e. TI-RADS), and cannot specifically draw conclusions about our models’ performance in comparison to these RSS in our patient population. Finally, overfitting of the model may be evidenced by the poor performance of the “All Predictors” model. This is likely due to the high number of predictors and the low number of observations on which to train and test the models. Overfitting appears to be introduced by the radiomics features in our analysis, because they are not particularly informative without a large sample size, so the fitted coeffcients for these predictors capture mostly noise rather than genuine statistics of image contrast that correlate with follicular adenoma/carcinoma.

## Conclusion

By focusing on contiguous dark and light regions, i.e. topological patterns, TDA can capture features from data that are potentially missed by less interpretable methods of imaging analysis. This novel methodology may augment current clinical decision-support tools for thyroid US interpretation. And importantly, advances in this field may assist with reducing the significant variability in interobserver interpretation of thyroid ultrasounds and may improve the standardization of thyroid nodule management.

## Data Availability

Data is not available for this study due to the small sample size to protect patient privacy.

