## Supplementary material for "A proof-of-concept investigation into predicting follicular carcinoma on ultrasound using topological data analysis and radiomics"

**Persistent homology**

Two-dimensional images only contain 0- and 1-dimensional topological features. To calculate the 0- or 1-dimensional persistent homology for a greyscale image $X$, we first require a *filtration function* $I$, on the image $X$. We then examine the shape of the binary images $I_t$, which consist of all pixels $p$ in $X$ that have pixel values $I(p)$ at most $t$. In what is called the *greyscale filtration* of the image, we range $t$ across all pixel values 0 to 255, take $I(p)$ to be the grayscale value of pixel $p$, and keep track of how and at which pixel values connected components (contiguous dark regions) and loops (contiguous white regions) appear. The value at which a given feature appears is called its *birth* value and the value at which it merges (or closes up) is called its *death* value. The collection of birth and death values of all the features within the 0- or 1-dimensional persistent homology of $I$ is called its (0- or 1-dimensional) *persistence diagram*.

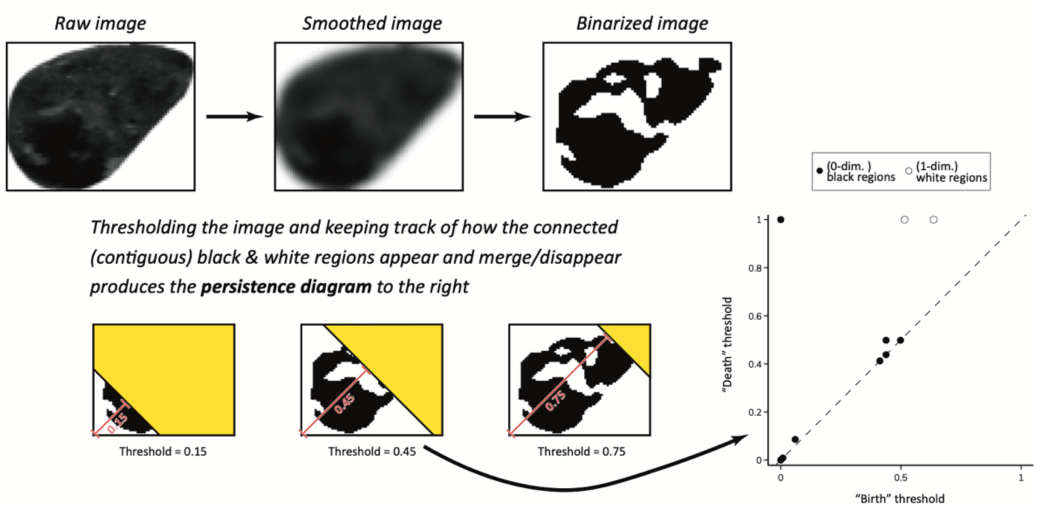


**Figure S1:** An illustration of the height filtration and its derived persistence diagram(s) for an ultrasound image of a follicular adenoma. The 180 geometric topological features were extracted using this method.

Recent work has established the utility of incorporating multiple types of topological and geometric information derived from persistence diagrams of various filtrations of images to improve predictive capabilities of machine learning models [1]. Two such filtrations are the height [1,2] and greyscale [3-6] filtrations. In the case of the height filtration an image is first binarized at some fixed threshold and the evolution of shape is examined as the black pixels are unveiled along on a given direction in the image—see **Figure S1**. The height filtration is motivated by theoretical results that establish a two- or three-dimensional shape is completely characterized by the persistence diagrams of its height filtrations [7]. Expanding upon this result, it was established that for a 2-dimensional binary image only 4 directions are needed [3].

As persistence diagrams are not immediately amenable to statistically analysis, we converted each diagram into a single numerical value via the persistent entropy [8]. Persistent entropy roughly captures how much shape information is associated to an image, with higher values indicating more shape complexity.

**<u>References</u>**


1. Garin A, Tauzin G. A Topological "Reading" Lesson: Classification of MNIST using TDA. 2019 18th IEEE International Conference On Machine Learning And Applications (ICMLA). 2019:1551-1556.
2. Oner D, Garin A, Koziński M, Hess K, Fua P. Persistent Homology with Improved Locality Information for more Effective Delineation. IEEE Transactions on Pattern Analysis and Machine Intelligence 2023. DOI:10.1109/TPAMI.2023.3246921.1-8.
3. Betthauser LM. Topological Reconstruction of Grayscale Images [Ph.D.]. United States -- Florida: University of Florida, 2018.
4. Chung YM, Hu CS, Lawson A, Smyth C. Topological approaches to skin disease image analysis. 2018 IEEE International Conference on Big Data (Big Data). 2018:100-105.
5. Rieck B, Yates T, Bock C, Borgwardt K, Wolf G, Turk-Browne N, et al. Uncovering the topology of time-varying fMRI data using cubical persistence. Advances in neural information processing systems 2020;33:6900-6912.

6. Thomas AM, Crozier PA, Xu Y, Matteson DS. Feature Detection and Hypothesis Testing for Extremely Noisy Nanoparticle Images using Topological Data Analysis. Technometrics 2023. DOI:10.1080/00401706.2023.2203744.1-14.
7. Turner K, Mukherjee S, Boyer DM. Persistent homology transform for modeling shapes and surfaces. Information and Inference: A Journal of the IMA 2014;3:310-344.
8. Rucco M, Castiglione F, Merelli E, Pettini M. Characterisation of the idiotypic immune network through persistent entropy. Proceedings of ECCS 2014: European conference on complex systems. Springer, 2016:117-128.
